# Genome-Wide Association Study of Coronary Microvascular Disease assessed by cardiac perfusion Positron Emission Tomography converges on NF-кB pathway

**DOI:** 10.1101/2025.03.20.25324357

**Authors:** Rasika Venkatesh, Tess Cherlin, Nicole Wayne, Rachit Kumar, Lindsay Guare, Venkata Singamneni, Brett Irving, Scott Dudek, Regeneron Genetics Center, Penn Medicine Biobank, Michael G. Levin, Shefali Setia-Verma, Marie A. Guerraty

## Abstract

**Background:** Coronary Microvascular Disease (CMVD) contributes to the large burden of ischemic heart disease, and there is a need for mechanistic insight and targeted therapies.

Perfusion cardiac PET allows for the quantitative assessment of myocardial blood flow reserve (MBFR), which reflects coronary microvascular function. Here we perform the first genome-wide association study (GWAS) using cardiac PET MBFR as a measure of CMVD.

**Methods:** MBFR was measured using Rubidium-82 cardiac perfusion PET obtained as part of routine clinical care. GWAS was performed using datasets from individuals genetically similar to EUR and AFR populations within the Penn Medicine Biobank (PMBB), followed by comprehensive downstream analyses including fine-mapping and transcriptome-wide association study (TWAS). We used gene set enrichment analysis (GSEA) to investigate associated molecular pathways and TIMI Frame Count to validate the association between the identified variants and CMVD. Finally, we assessed associations between key loci and proteins in the NF-**к**B pathway in the UK Biobank Pharma Proteome pGWAS dataset and with individual-level OLINK proteomic data in PMBB.

**Results:** Among 241 targeted CAD loci, 17 in the AFR, 14 in the EUR, and 16 shared across both populations were significant (p<2e-04). A subsequent discovery GWAS identified 1 genome-wide significant association. Fine mapping identified a signal near *ERC1*. Sex-stratified TWAS identified C*APN2* in females and *PLA2G5* in males. GSEA identified inflammatory and NF-**к**B pathways as the top pathways associated with these loci, and TIMI frame count confirmed that these loci were protective. Using proteomic data, we found IL-1B was increased in individuals with CAPN2 variant, and NEMO was differentially regulated by CAPN2 and ERC1 variants.

**Conclusions:** Our study identified variants associated with MBFR in populations of EUR and AFR ancestry and converged on two independent loci that are near genes known to regulate the NF-**к**B pathway. Our multi-omic analyses support a role for NF-**к**B pathway in CMVD.

## Introduction

Coronary microvascular disease (CMVD), defined as disease of the coronary pre-arterioles, arterioles, and capillaries, accounts for 30-50% of the burden of ischemic heart disease.^1–4^ However, our understanding of CMVD remains limited, and there are no targeted therapies for CMVD.^5^ Cardiac perfusion positron emission tomography (PET) has emerged as a leading non-invasive imaging modality to assess for CMVD. Myocardial blood flow reserve (MBFR) assessed by PET is the ratio of MBF under hyperemic conditions to rest MBF and represents the ability of the coronary microcirculation to vasodilate appropriately, such that MBFR < 2 is considered CMVD.^6^

Recent genome-wide association studies (GWAS) of coronary artery disease (CAD), encompassing data from hundreds of thousands of patients, have identified over 241 genetic loci associated with CAD.^7–9^ However, inclusion criteria are often broad, allowing for the inclusion of patients with heterogeneous phenotypes. For instance, in Shunckter et al, participants were included on the basis of either confirmed obstructive CAD through coronary angiography or based on electrocardiogram (ECG) changes, symptoms, and serologic evidence of myocardial damage without definitive confirmation of CAD.^9^ Such criteria overlap with those for CMVD, or ischemia with no obstructive coronary artery disease (INOCA); as such, it is probable that a significant proportion of these patients may have CMVD as the underlying cause of their ischemic symptoms. We, therefore, postulate that existing CAD GWAS might more aptly be described as ischemic heart disease (IHD) GWAS and include genetic associations with both CAD and CMVD.^10^ By leveraging these existing “IHD loci”, we can refine our genetic interrogation, potentially enhancing statistical power and precision.

In this study, we investigate the genetics of CMVD with data from the Penn Medicine BioBank (PMBB) using myocardial blood flow reserve (MBFR) measured by perfusion PET as the phenotype for CMVD. We first performed a targeted association study of 241 previously identified “IHD loci.” We then perform a new GWAS for MBFR in participants who were genetically similar to African (AFR) and European (EUR) populations from the PMBB cohort. To better understand the molecular underpinnings of CMVD, we used downstream analyses, stratified by sex, to identify an association between CMVD and the NF-**к**B pathway. We then used thrombolysis in myocardial infarction (TIMI) frame count to phenotypically validate the variants. Finally, we further link the NF-**к**B pathway and the molecular pathophysiology of CMVD with using targeted proteomics.

With the convergence of advanced imaging techniques, robust genetic data, multi-omics pathway analyses, and clinical validation, this study offers novel insights into the genetic underpinnings of CMVD.

## Methods

### Study Population

The Penn Medicine Biobank (PMBB) represents a relatively diverse population (mean age 56.6 ± 15.1 yrs; 55.52% male; 75.46% European, 18.7% African American, 2.09% Asian; 1.75% Hispanic). 43,625 participants have been genotyped and imputed using the TOPMed reference panel; each participant’s genetic data is linked with their Electronic Health Records (EHR),^11^ including perfusion PET stress testing data. The study was approved by the University of Pennsylvania Institutional Review Board, and all participants consented to participate. Principal component analyses were performed on all samples and genetically informed ancestry was determined by projecting PMBB participants on the 1000 Genomes Project refence population.^12^ Age at the time of exam, Body Mass Index (BMI), and sex were extracted from the EHR. We additionally extracted ICD codes reflective of comorbidities of CAD and CMVD. Schematic of cohorts and datasets used are summarized in Supplemental Figure 1.

### PennMedicine Biobank Perfusion PET GWAS Cohort

We included 1,002 patients with available genotyping from PMBB who had undergone Rubidium-82 (Rb-82) perfusion PET stress testing at the Hospital of the University of Pennsylvania between 2012 and 2020 as part of routine clinical care and enrolled in PMBB. Patients with incomplete or poor-quality myocardial blood flow reserve data were excluded (n=80). The remaining 383 participants of European (EUR) ancestry and 539 participants of African (AFR) ancestry were analyzed for genomic association. All patients received dipyridamole or regadenoson for coronary vasodilation, and imaging data was analyzed using Siemens Syngo MBF (prior to 2/1/2020) or Invia Corridor 4DM (after 2/1/2020). Subsequent analyses were adjusted for analysis software. Plasma samples for 879 of these patients were available through PMBB and underwent proteomic profiling using OLINK’s Cardiovascular II panel.

### PennMedicine Biobank Angiography Cohort

To orthogonally validate associations between variants rs115240131 (ERC1) and rs6700366 (CAPN2) and CMVD, we compared the TIMI frame count (TFC) for patients in PMBB with and without the variants.^13^ Briefly, a blinded reviewer quantified corrected TFC in unobstructed left anterior descending (LAD) coronaries in 2 cohorts of PennMedicine Biobank participants matched for ERC1 (n = 123) and CAPN2 (n = 95) where each participant was matched 1:20:20 for two copies of the alternate allele (ALT) (n = 3), heterozygosity (HET), and reference (REF) for ERC1 loci and 1:2:2 for ALT (n = 19), HET, and REF for CAPN2 loci, respectively. Data are presented as mean±SD, and T-test was used to compare groups. Coronary angiographs obtained as part of routine clinical care obtained at 15 frames per second were used, and the final TFC measurement was multiplied by 2 to standardize to angiography images used by Gibson et al., which were acquired at a frame rate of 30 frames per second.^14^ For all patients with TFC measurements, coronary angiography images were used to determine the presence or absence of obstructive CAD, defined as coronary obstruction greater than or equal to 50% in one or more coronary artery. The rate of obstructive CAD was compared between patients with and without the variant. Percentage of patients in each group with obstructive CAD were compared using chi-squared test.

### Genomic Association Analyses

Our primary objective was to discern genetic loci associated with coronary microvascular disease (CMVD) using a genome-wide association study (GWAS) approach. As an initial step, we conducted a targeted association study between known CAD loci and MBFR using REGENIE and adjusted the analyses by PCs. Subsequently, we performed a comprehensive GWAS stratified by sex and genetically inferred ancestry using REGENIE.^15^ To ensure robustness and account for potential confounders, our models were adjusted for various parameters, including age, sex, age squared, interaction between age and sex, the first five PCs, and the batch corresponding to phenotype extraction. The results from the ancestry stratified analyses were then synthesized in a meta-analysis using PLINK 2.0,^16^ providing a consolidated view of the genetic landscape associated with CMVD.

### Fine-Mapping of GWAS Loci

Fine mapping was performed using PolyFun,^17^ a SuSiE-based fine-mapping approach that incorporates functional annotations, for loci with p-value < 1e-05 from the CMVD GWAS. As GWAS considers associations only in tagged variants, fine mapping can identify the most functionally informed variants in these regions with strong associations to the MBFR phenotype. To enhance the precision of our mapping, we incorporated gene expression data and functional annotations from various cardiovascular tissues, namely the heart ventricles, atrium, aorta, and coronary arteries, sourced from the GTEx database. We further included annotations of nonregulatory regions, such as enhancers and promoters, using the EpiMap database.^16^

### Transcriptome-Wide Association Study (TWAS)

We subsequently performed tissue-specific transcriptome-wide association studies (TWAS) using S<u>-</u>PrediXcan with the GWAS summary statistics from our GWAS analyses.^18^ TWAS was performed on AFR and EUR samples separately, with and without sex stratification; three TWAS (female only, male only, and both sexes) were ultimately performed for each ancestry grouping using different eQTLs. One eQTL set consisted of heart failure eQTLs derived from 136 nonfailing donor heart samples within the MAGNET consortium at a nominal p-value of 0.001. Models for two heart tissues were sourced from PredictDB; these models make use of GTEx gene expression data in two relevant tissues: heart left ventricle and heart atrial appendage. S-PrediXCan TWAS models were constructed using MAGNET consortium data by utilizing PEERS covariates, expression information from eQTL associations, gene and SNP annotations.^19^ Existing mashr GTEx models were also incorporated for the heart left ventricle and heart atrial appendage tissues.^20^ The resulting TWAS associations were assessed for statistical significance using a Benjamini-Hochberg (FDR) threshold of 0.05 for each ancestry group, after stratification by sex.

### Pathway analyses using MAGNET data

To understand how the top variants identified in our analyses affected gene expression in cardiac tissue, we performed differential gene expression analyses and gene set enrichment analyses using the MAGNET data.^21^ For this analysis, we used non-failing control heart expression data and included male and female samples from EUR ancestry (n=122) since this afforded increased power. For rs839696, rs6700366, and rs115240131 (in the genes *ADK* (negative control), *CAPN2*, and *ERC1* respectively), we performed differential gene expression analysis between tissues from participants heterozygous for the alternative allele relative to reference samples using the DEseq2 algorithm as implemented in Partek Flow (version 11.0.23.1105). We then performed gene set enrichment analysis using hallmark pathways.^22^

### UK Biobank pGWAS Analysis

We then analyzed summary statistics from a proteome-wide GWAS performed using circulating plasma protein quantitative trait loci (pQTLs) from the UK Biobank Pharma Proteome Study.^23^ We measured the association of variants from our genes of interest (*ADK, CAPN2, ERC1, PLA2G5*, and *AADAC*) with the expression of selected proteins (CXCL10, CXCL11, IL-1B, and IL-6) from the NFKB pathway for the available Inflammation, Neurology, and Cardiometabolic phenotypes. The summary statistic results were compared across the Combined, African, and European discovery cohorts at a Benjamini-Hochberg (FDR) significance threshold of 0.05 after p-value adjustment.

## Results

The study cohort consisted of 1,002 patient participants. Of these, 515 were males and 487 were females. Genetic analyses were employed to determine ancestry, with 573 and 429 samples being most genetically similar to African (AFR) and European (EUR) reference populations, respectively. The cohort had high prevalence of cardiac co-morbidities, including hypertension, obesity, and type 2 diabetes (T2D) as shown in Table 1. The mean examination age of the participants was 60.6 years, and the average BMI for the cohort was recorded at 34.9 kg/m2. Due to the high prevalence of cardiovascular risk factors, the mean MBFR observed for the population is close to 2, reflecting a similar number of participants with and without CMVD. Participants of African ancestry had higher MBFR relative to those of European ancestry, consistent with previously reported observations.^6^

**Table 1:**
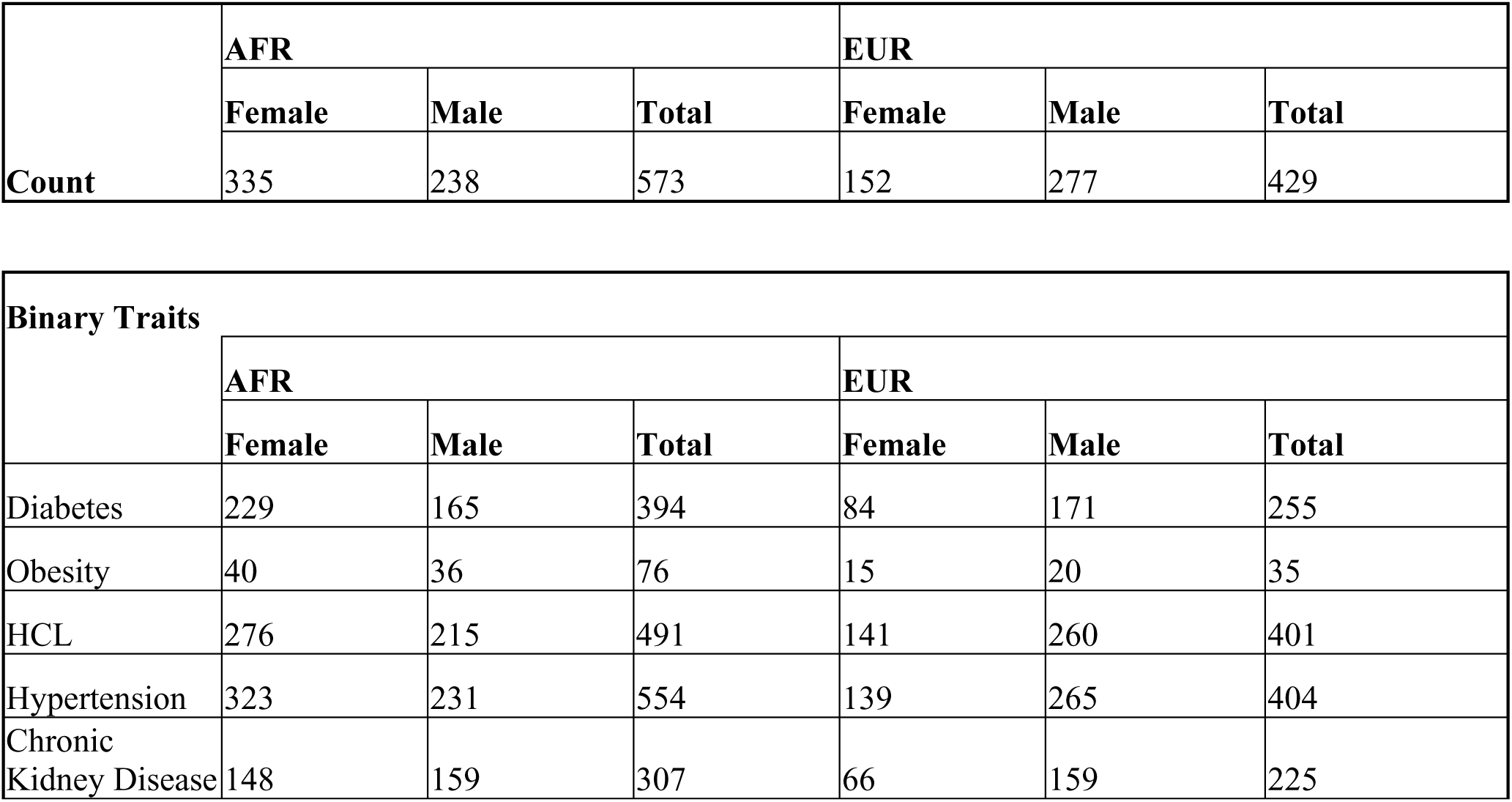
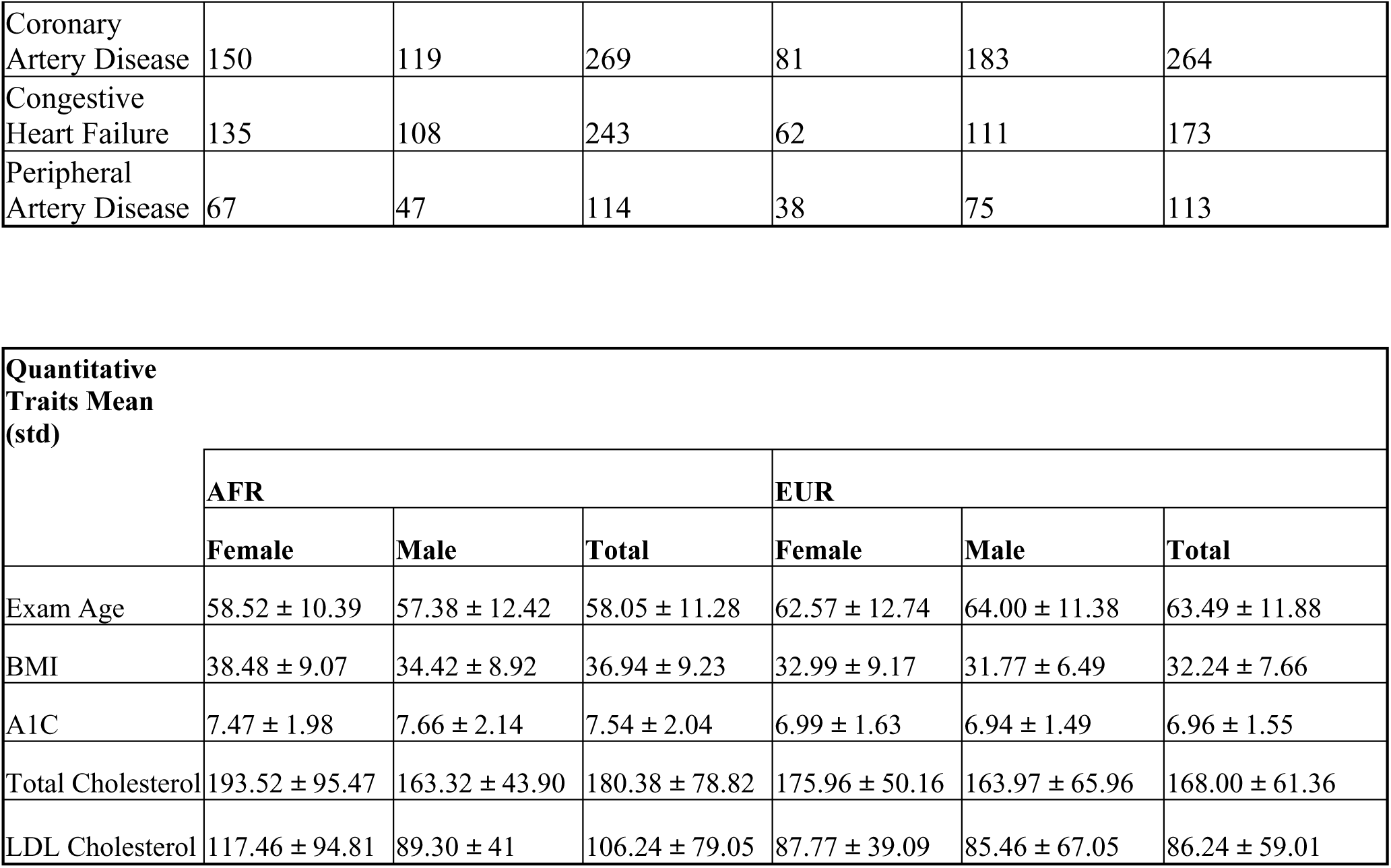
Demographics and summary of Penn Medicine BioBank participants included in GWAS.

### Association of Known “IHD Loci” with MBFR

We conducted an association analysis between MBFR as assessed by perfusion PET and previously identified IHD GWAS loci. Out of the 241 distinct loci tested, 17 were significant in the AFR population, 14 in the EUR population, and 16 in both analyses at the Bonferroni threshold (p≤0.0002). These results are reported in Supplemental Table 1 and also highlighted in Figure 1. Notably, several significant loci are known to play a role in microvascular health or angiogenesis, including *FLT1, KLF4, NOS3, ITGA1*, *SMAD3, PPAP2B, DNAH10 and PRDM16* (Table 2). For example, FLT1 is the main VEGFA receptor and regulates endothelial angiogenesis and microvascular stability;^24^ KLF4 is necessary for the maintenance of coronary microvascular pericyte coverage;^25^ and NOS3 expression is crucial for proper coronary microvascular function.^26^ Of note, the beta values for these key genes is consistent with those seen in CAD analyses, reinforcing the concept that some loci associated with myocardial infarction or angina to be due to CMVD instead of or in addition to CAD.

**Figure 1.**
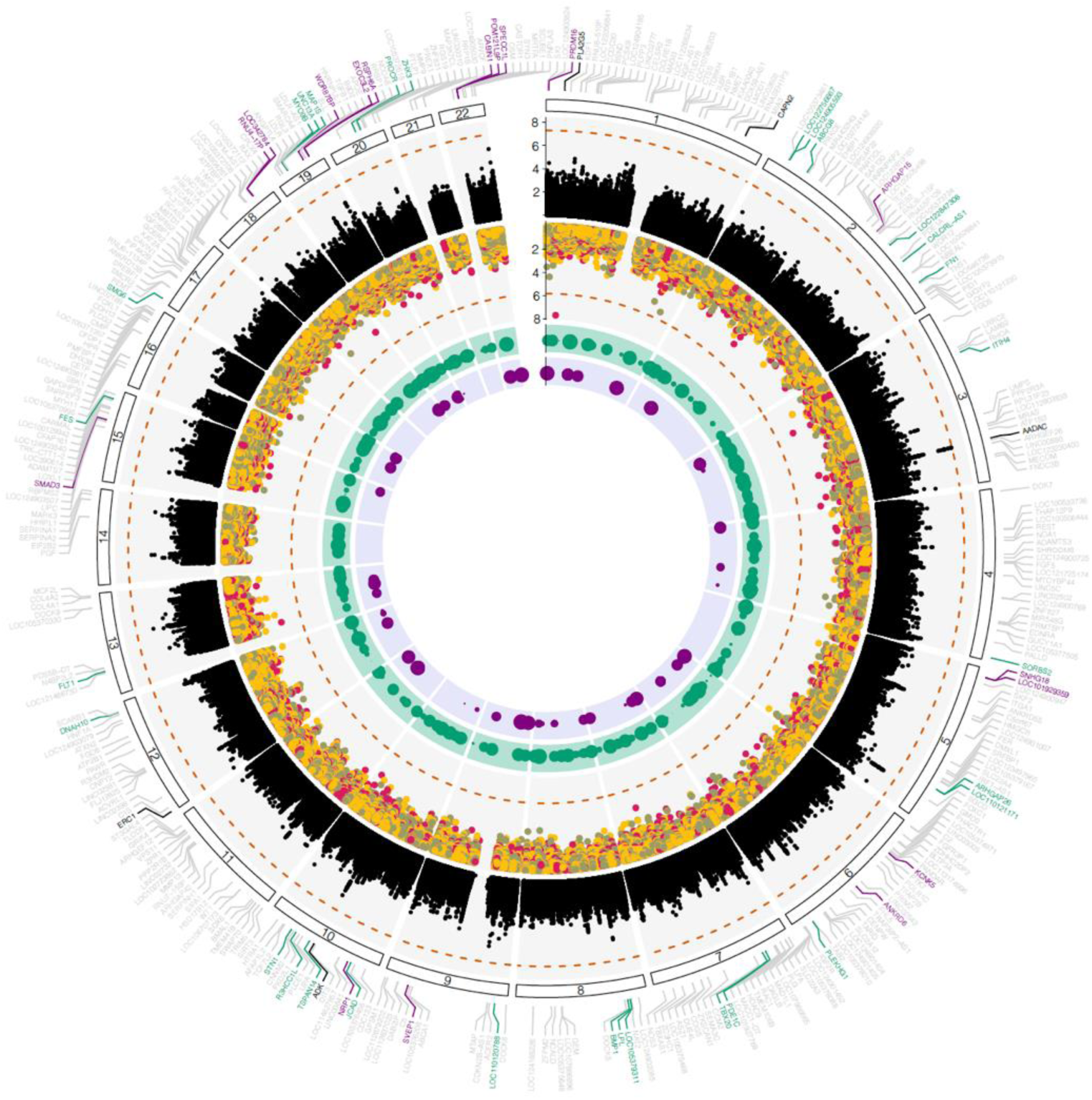
**Summary of GWAS and downstream analyses**. Circos Plot of Meta-Analyses and annotations for CAD Loci. A multi-layered circos plot, provides a holistic view of the genetic landscape associated with CMVD assessed by MBFR. The outermost layers is showing annotations across all autosomes for the 241 known CAD loci. Loci that replicated in AFR participants cohort are highlighted in green, while those specific to EUR participants are depicted in purple. Noteworthy loci near *AADAC*, *CAPN2*, *ERC1*, and *PLA2G5* are indicated in black.

**Table 2:** Subset of 241 IHD loci associated with MBFR in AFR, EUR, and ancestry meta-analysis CMVD GWAS results.

|  | Myocardial Blood Flow Reserve |  |  |  |  |  |  |
| --- | --- | --- | --- | --- | --- | --- | --- |
| NEAREST GENE | RSID | A1 | A2 | BETA | P | SE | POP |
| PRDM16 | rs10909950 | G | C | 0.182143 | 4.71e-06 | 0.039792612 | EUR |
| PRDM16 | rs4648398 | C | T | -0.159263 | 2.61e-05 | 0.037874032 | AFR |
| PRDM16 | rs10909950 | G | C | 0.182143 | 4.71e-06 | 0.039792612 | EUR |
| PRDM16 | rs4648398 | T | C | -0.126243727 | 3.17e-05 | 0.030340507 | META |
| PPAP2B | rs6656021 | A | T | 0.200898 | 0.000131 | 0.052528245 | EUR |
| PPAP2B | rs6656021 | A | T | 0.200898 | 0.000131 | 0.052528245 | EUR |
| CELSR2 | rs111755757 | G | T | 0.23285623 | 2.84e-05 | 0.055628305 | META |
| APOB | rs114863832 | A | G | 0.404636 | 5.4e-05 | 0.100216871 | AFR |
| ABCG8 | rs115066330 | A | C | 0.324292 | 8.13e-05 | 0.082296434 | AFR |
| ABCG8 | rs115066330 | A | C | 0.324292 | 8.13e-05 | 0.082296434 | AFR |
| PRKCE | rs4952798 | G | A | -0.118333187 | 0.00019 | 0.031708102 | META |
| ARHGAP15 | NA | A | AT | 0.172693 | 0.000116 | 0.044803782 | EUR |
| ARHGAP15 | rs28396876 | G | A | 0.121952111 | 0.000173 | 0.032472989 | META |
| ITIH4 | rs147348595 | G | T | 0.44993 | 9.27e-05 | 0.115102663 | AFR |
| FNDC3B | rs7611863 | T | C | 0.166530871 | 9.32e-05 | 0.042616681 | META |
| SNORD123 | rs12520881 | T | C | 0.192193 | 1.08e-05 | 0.04367523 | EUR |
| SNORD123 | rs12520881 | C | T | 0.143234168 | 7.23e-05 | 0.03609198 | META |
| ITGA1 | rs830866 | G | A | 0.158199615 | 0.000127 | 0.041281619 | META |
| ARHGAP26 | rs17099100 | A | G | 0.179165 | 3.12e-05 | 0.043021702 | AFR |
| ARHGAP26 | rs17099100 | G | A | 0.157858085 | 5.41e-05 | 0.039101179 | META |
| KCNK5 | rs2206860 | T | G | 0.138573 | 0.000117 | 0.035971233 | EUR |
| KCNK5 | rs2206860 | G | T | 0.103909463 | 7.31e-05 | 0.026200306 | META |
| SLC22A1 | rs755883 | C | G | 0.157267 | 0.000179 | 0.041972039 | AFR |
| TBX20 | rs6977729 | A | G | -0.12397718 | 8.63e-05 | 0.031576924 | META |
| NOS3 | rs73480177 | C | G | 0.146953413 | 8.33e-05 | 0.037348042 | META |
| LPL | rs59187853 | C | A | 0.27695 | 7.2e-05 | 0.069768119 | AFR |
| LPL | rs59187853 | C | A | 0.27695 | 7.2e-05 | 0.069768119 | AFR |
| SLC18A1 | rs59187853 | C | A | 0.27695 | 7.2e-05 | 0.069768119 | AFR |
| BMP1 | rs13276612 | T | C | 0.153085 | 6.2e-05 | 0.038222835 | AFR |
| BMP1 | rs13276612 | C | T | 0.104450101 | 0.000134 | 0.0273502 | META |
| C9orf146 | rs80248213 | A | G | 0.426395 | 5.07e-05 | 0.105220975 | AFR |
| KLF4 | rs260223 | A | G | 0.184261 | 8.36e-05 | 0.046840002 | EUR |
| KLF4 | rs4574919 | G | T | 0.103097958 | 8.75e-05 | 0.026281251 | META |
| TRIM5 | rs2736575 | A | G | 0.10625012 | 7.24e-05 | 0.026775005 | META |
| DNAH10 | rs1242999 | A | G | 0.167466 | 0.000166 | 0.044469936 | EUR |
| SCARB1 | rs1992892 | A | G | 0.109392377 | 0.000124 | 0.028501848 | META |
| SCARB1 | rs1992892 | A | G | 0.109392377 | 0.000124 | 0.028501848 | META |
| FLT1 | rs7989324 | T | C | -0.154665 | 5.09e-05 | 0.038175169 | AFR |
| SMAD3 | rs116870050 | G | A | 0.355865 | 5.4e-05 | 0.088137677 | EUR |
| CMIP | rs62044094 | T | C | 0.113328685 | 6.4e-05 | 0.028349497 | META |
| PLCG2 | rs62044094 | T | C | 0.113328685 | 6.4e-05 | 0.028349497 | META |
| CDH13 | rs62035221 | A | G | 0.280378 | 0.000165 | 0.074423446 | AFR |
| MYO9B | rs78019026 | G | A | 0.513361 | 3.96e-06 | 0.111274359 | AFR |
| MYO9B | rs4808061 | A | G | -0.171342 | 4.33e-05 | 0.041901855 | EUR |
| MAP1S | rs78019026 | G | A | 0.513361 | 3.96e-06 | 0.111274359 | AFR |
| APOE | chr19:45571319 | ATT | A | 0.151292 | 0.000111 | 0.039142128 | EUR |
| APOE | chr19:45571319 | ATT | A | 0.151292 | 0.000111 | 0.039142128 | EUR |
| EXOC3L2 | chr19:45571319 | ATT | A | 0.151292 | 0.000111 | 0.039142128 | EUR |
| ARVCF | rs5748310 | G | A | -0.217139 | 0.000177 | 0.057907354 | AFR |
| CABIN1 | rs200148140 | A | AT | 0.182902 | 7.11e-05 | 0.046041176 | AFR |
| CABIN1 | rs1402156941 | GCC | G | 0.182697 | 5.94e-05 | 0.045501552 | EUR |
| SPECC1L | rs200148140 | A | AT | 0.182902 | 7.11e-05 | 0.046041176 | AFR |
| SPECC1L | rs1402156941 | GCC | G | 0.182697 | 5.94e-05 | 0.045501552 | EUR |

### Genome-wide association analyses of MBFR

We next extended our analysis and performed a discovery GWAS for participants of both African (AFR) and European (EUR) genetic ancestries from PMBB. Meta-analysis of these GWAS results resulted in one locus rs1969348, that approached GWAS significance (p-value< 5×10^-8^) as shown in Figure 1. All results stratified by ancestry are represented in Supplemental Tables 2, 3, and 4. The variant rs1969348 mapped to the *AADAC* gene (chromosome 3). *AADAC* has been shown to regulate smooth muscle cell migration and proliferation and protect against atherosclerosis.^27^ A variant near *ADK* (rs839696 on chromosome 1) was near-significantly associated with MBFR (p-value = 1.17×10^-6^). Perfusion PET stress testing relies on an adenosine analog to induce hyperemia, and individuals with altered ADK may have an altered response to the stress test. Alternately, *ADK* gene has been shown to play a role in cardiomyocyte microtubule dynamics and is protective against cardiac hypertrophy.^28^ Finally, variants in the *TIMD4* gene, another near-significant loci associated with MBFR (p = 8.59E-07), has been shown to play a role in lipid regulation and atherosclerosis.^29, 30^

A circos plot in Figure 1 offers a comprehensive visualization of the meta-analysis results, known “IHD loci”, and additional layers of genetic and transcriptomic data from post-GWAS analyses to help elucidate the genetic underpinnings of CMVD.

The second layer displays the Manhattan plot from the meta-analyses of CMVD GWAS, with significant associations represented above the red dotted line (p = 5 x 10^-8^). Moving inward, the next circle displays the Transcriptome-wide association study (TWAS) results. Points within this circle are color-coded based on the source of gene expression data: GTEx-derived results from heart atrium are in yellow, heart left ventricle in military green while those from the MAGNet consortium are shown in magenta. The innermost circles represent the results of sex-based heterogeneity analyses. The I˄2 statistic, indicative of the proportion of total variation in study estimates due to heterogeneity, is visualized in two separate circles. The green circle corresponds to the AFR cohort, and the purple circle represents the EUR cohort.

### Fine-mapping analysis

To gain insight into credible gene sets which may be associated with MBFR, we performed fine-mapping analysis using EpiMap and GTEx data from Heart Atrial Appendage and Heart Left Ventricle tissues. The full results of fine-mapping are reported in Supplemental Table 5. At a posterior inclusion probability (PIP) threshold > 0.75, we identified rs115240131 in *ERC1 gene* and rs4491579 in *LOC105371755.* An additional association in gene *NDUFA5* (rs116069955) was identified as approaching significance (PIP = 0.749). Interestingly, SNPs that map to ERC1 have previously been associated with spontaneous coronary dissection.^31^

### Transcriptome Wide Association (TWAS) analysis

We next performed ancestry and sex-stratified transcriptome-wide association study (TWAS). Results of the TWAS can be found in Supplemental Table 6 and the third track of the circos plot in Figure 1. One significant association with the MBFR phenotype emerged from the AFR cohort analyses as well as one association approaching significance after Benjamini-Hochberg (FDR) correction (p-value< 0.05). Specifically, in the AFR female cohort, *CAPN2* approached transcriptome-wide significance (uncorrected p-value=6.9e-6) when using eQTLs obtained from the mashr Heart Left Ventricle model in the GTEx v8 database. Additionally, *PLA2G5* was identified as significantly associated with the MBFR phenotype in the AFR Male cohort (uncorrected p-value =2.2e-8) in the model trained on heart failure data from the MAGNet consortium. The full results across ancestries and sexes with FDR correction are reported in Supplemental Table 6.

### ERC1 and CAPN2 loci are associated with protection against CMVD

Analyses downstream of GWAS identified rs115240131 (ERC1) and rs6700366 (CAPN2) as associated with CMVD. To determine the directionality and validate the association with a coronary microvascular phenotype, we examined individual level data for MBFR by genotype (Figure 2A). rs6700366 and rs115240131 were associated with increased MBFR suggestive of protection against CMVD.

**Figure 2.**
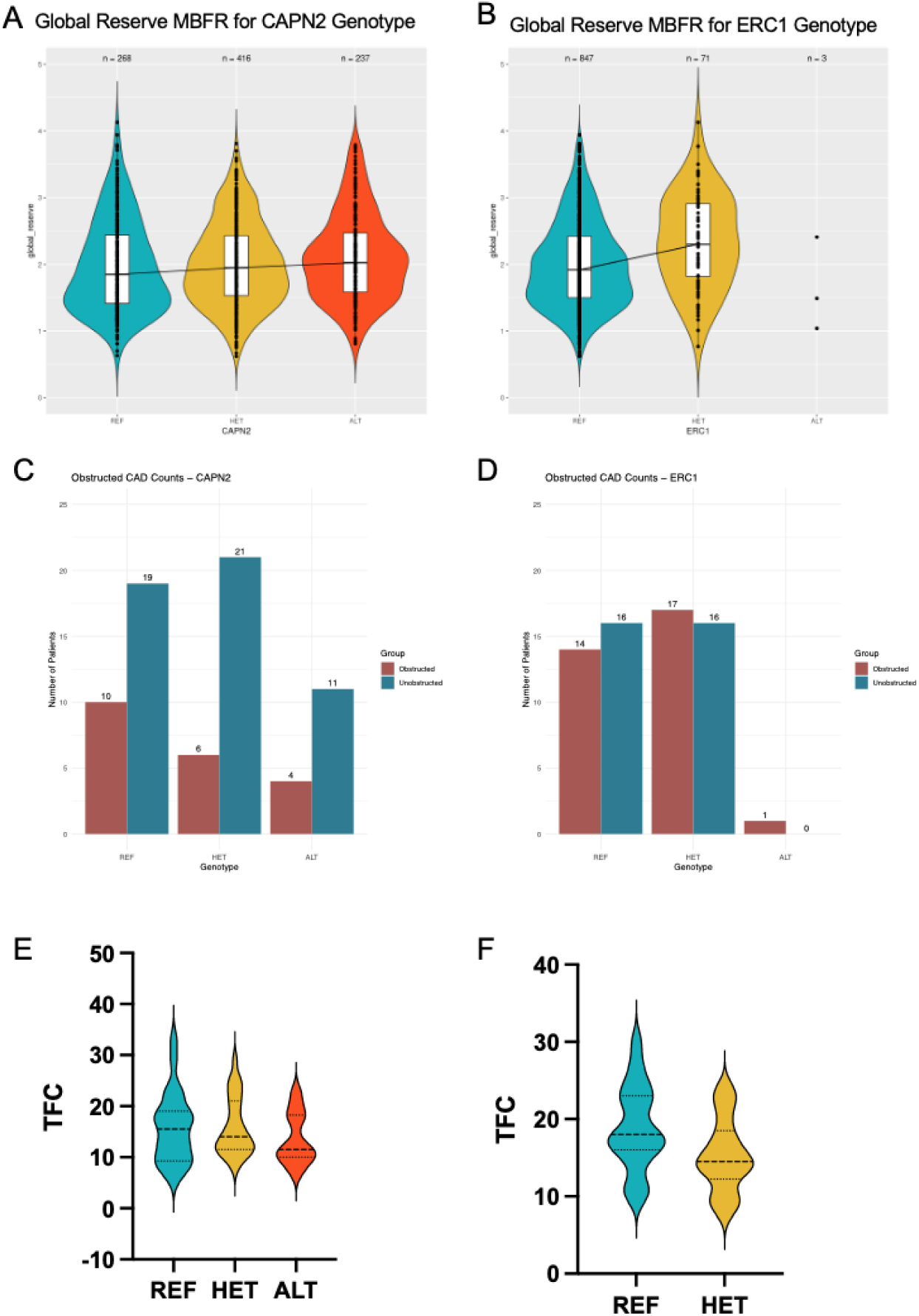
rs115240131 (ERC1) and rs6700366 (CAPN2) are consistent with protection against CMVD. (A,B) Violin plots of MBFR from individuals in the PMBB by genotype for REF (blue), HET (yellow), and ALT (red) alleles. Medians are connected across genotypes to show trends in MBFR phenotype. Number of counts for each genotype are indicated at the top of each plot. (C,D) No difference in obstructive CAD noted for individuals by genotype. (E,F) TIMI frame count was lower in individuals with rs115240131 (left) or rs6700366 (right) in a distinct cohort in PMBB.

We next sought to confirm whether these findings were indeed due to changes caused by CMVD rather than epicardial CAD. We manually reviewed coronary angiograms for patients with the variants identified near ERC1 and CAPN2 in a distinct cohort of individuals in PMBB (see Methods section). We found no difference in burden of obstructive CAD (defined as 1 or more coronary vessels with >=50% stenosis) between individuals with and without the variants in CAPN2 and ERC1 (Figure 2B). To orthogonally validate the association between the variants and CMVD, we used Thrombolysis in Myocardial Infarction (TIMI) frame count. TIMI frame count has been used to assess the coronary microvasculature, and we have previously shown that TFC correlates with presence of CMVD in patients without obstructive CAD and can serve as a measure of CMVD.^32^ We found that individuals with the ERC1 variant had lower TFC (e.g. brisker coronary flow) (22.1 +/-6.5 vs 18 +/-5.5, p<0.05, Figure 2E). Similarly, in a dominant negative model, there was a trend towards an association between CAPN2 variant and lower TFC (22.5 +/-8.1 vs 19.5 +/-6.8, p = 0.18, Figure 2F). These data support an association between rs6700366 (CAPN2) and rs115240131 (ERC1) and CMVD rather than CAD.

### Pathway analysis identifies a link between rs115240131 (ERC1) and rs6700366 (CAPN2) and *NF-**к**B pathway*

To further understand how the loci near ERC1 and CAPN2 could be protective against CMVD, as informed by the beta values above, and gain insight into potential mechanisms, we performed differential gene expression and gene set enrichment analyses using MAGNET data. For each variant, RNA-seq data from control, non-failing hearts (n=122) was compared between participants heterozygous for the respective variant relative to participants homozygous for the major allele. For rs115240131 (ERC1), there were 110 individuals with the AA (REF) genotype and 12 heterozygous individuals with AC genotype. Gene set enrichment analysis (GSEA) identified inflammatory and NF-**к**B pathways as the top pathways associated with both ERC1 and CAPN2 loci (Enrichment scores 4.42, p=0.01 and 3.94, p=0.02) (Figure 3A). Of note, a similar analysis using the loci identified near ADK did not identify NF-**к**B pathway as differentially regulated, serving as an important negative control (Supplemental Figure 2).

**Figure 3.**
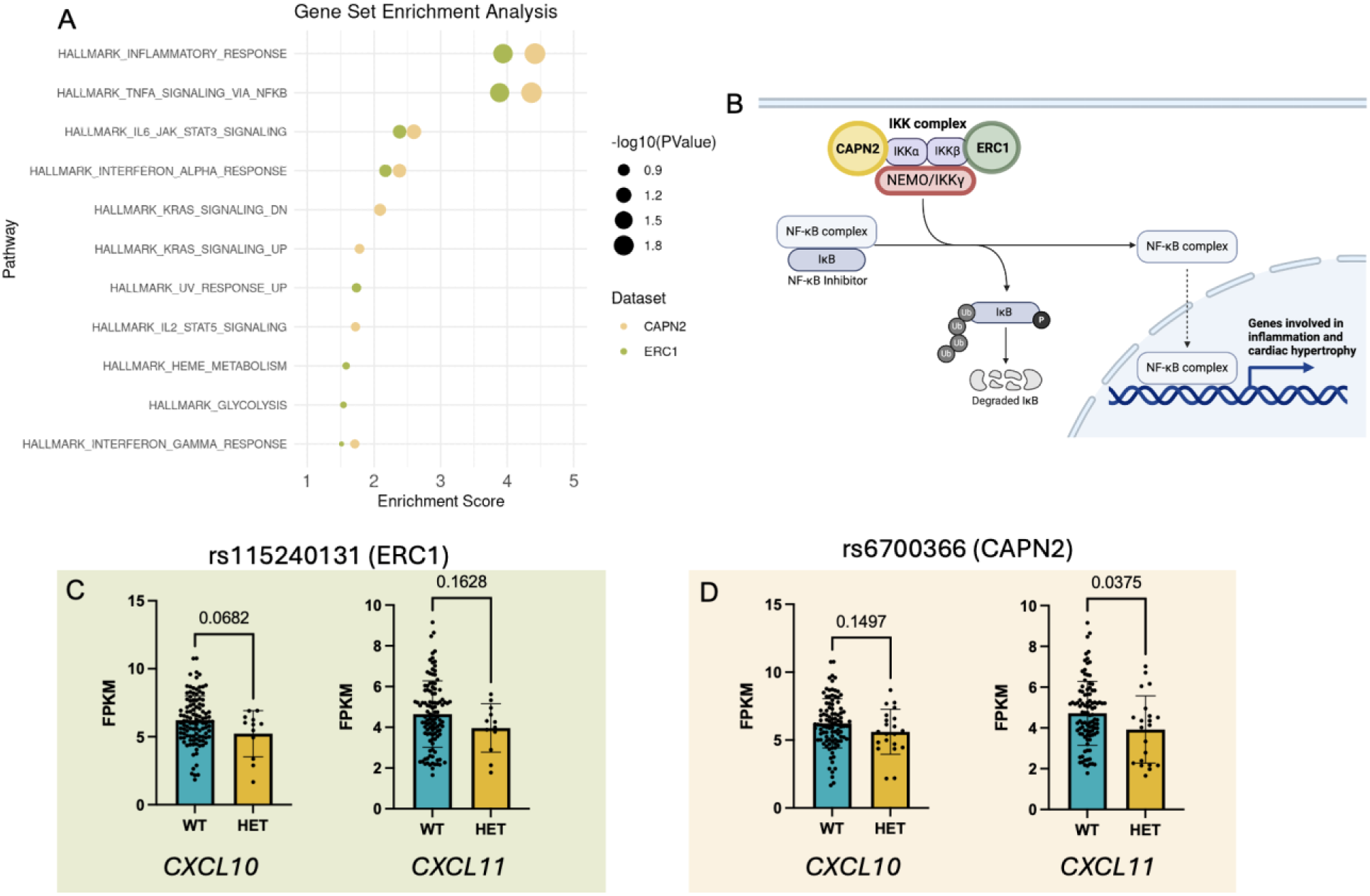
Gene and Pathway analysis for rs115240131 (ERC1) and rs6700366 (CAPN2). (A) Gene set enrichment analysis of genes differentially expressed between heart tissue from individuals with SNP and control by size of points (-log10(p-value)). (B) Schematic of NF-**к**B pathway with CAPN2 in light green and ERC1 in dark green. (C-D). Boxplots of showing gene expression of *CXCL10* and *CXCL11* is decreased in carriers of rs115240131 (C, green) or rs6700366 (D, yellow).

The NF-**к**B pathway is a master regulator of inflammation and plays important roles in angiogenesis and cardiac hypertrophy. NF-**к**B is usually bound to its inhibitor I**к**B, and remains inactive in the cytoplasm. Activation of the IKK complex leads to I**к**B degradation and nuclear translocation of NF-**к**B where it can increase target gene expression. *ERC1* is known to interact with IKKB and is required for NF-**к**B activation.^33^ Similarly, CAPN2 has been shown to interact with IKKa subunit of the IKK complex to promote NF-**к**B activation.^34^

The GSEA signal was driven largely by differential expression of CXCL10 (FPKM 6.2±1.7 vs 5.2±1.7, p=0.068) and CXCL11 (FPKM 4.7±1.6 vs 3.9±1.6, p<0.05) for control vs participants heterozygous for rs115240131 and rs6700366, respectively. These are known targets of NF-**к**B and act as inhibitors of angiogenesis (Figure 3C-D), such that a decrease in these factors would be protective. To determine which cell types may contribute to this result, we visualized ERC1 and CAPN2 expression in cardiac and coronary artery cell types using the tabula sapiens browser (Supplemental Figure 3).^35^ CAPN2 is most notably expressed in endothelial cells, with very low expression noted in immune cells, smooth muscle cells (SMCs), pericytes, and cardiomyocytes. ERC1 expression was higher in general when compared with CAPN2 with highest expression in endothelial cells and pericytes and moderate expression in immune cells, SMCs, and cardiomyocytes.

### Plasma proteomic data support an association between variants and inflammatory pathways

Given the association between rs115240131 (ERC1) and rs6700366 (CAPN2) and NF-**к**B pathway, we analyzed UK Biobank Pharma Proteome pGWAS summary statistics to determine whether these variants were associated with plasma protein levels of anti-angiogenic proteins CXCL10, CXCL11 and NF-**к**B-related inflammatory mediators IL-1B and IL6. We found no differences by genotype in CXCL10, CXCL11, or IL6. The heatmap in Figure 4A depicts the unadjusted –LOG10 P values of significant variants with an FDR-adjusted p-value of < 0.05, as well as their corresponding effect size (BETA). Notably, in the AFR cohort, *CAPN2* variants were found to be significant in association with plasma IL-1B levels. These findings were not noted in the EUR or combined ancestry cohorts from the UKBB Pharma Proteome pGWAS datasets, suggesting a more significant effect in individuals of African genetic ancestry (Supplemental Table 7).

**Figure 4.**
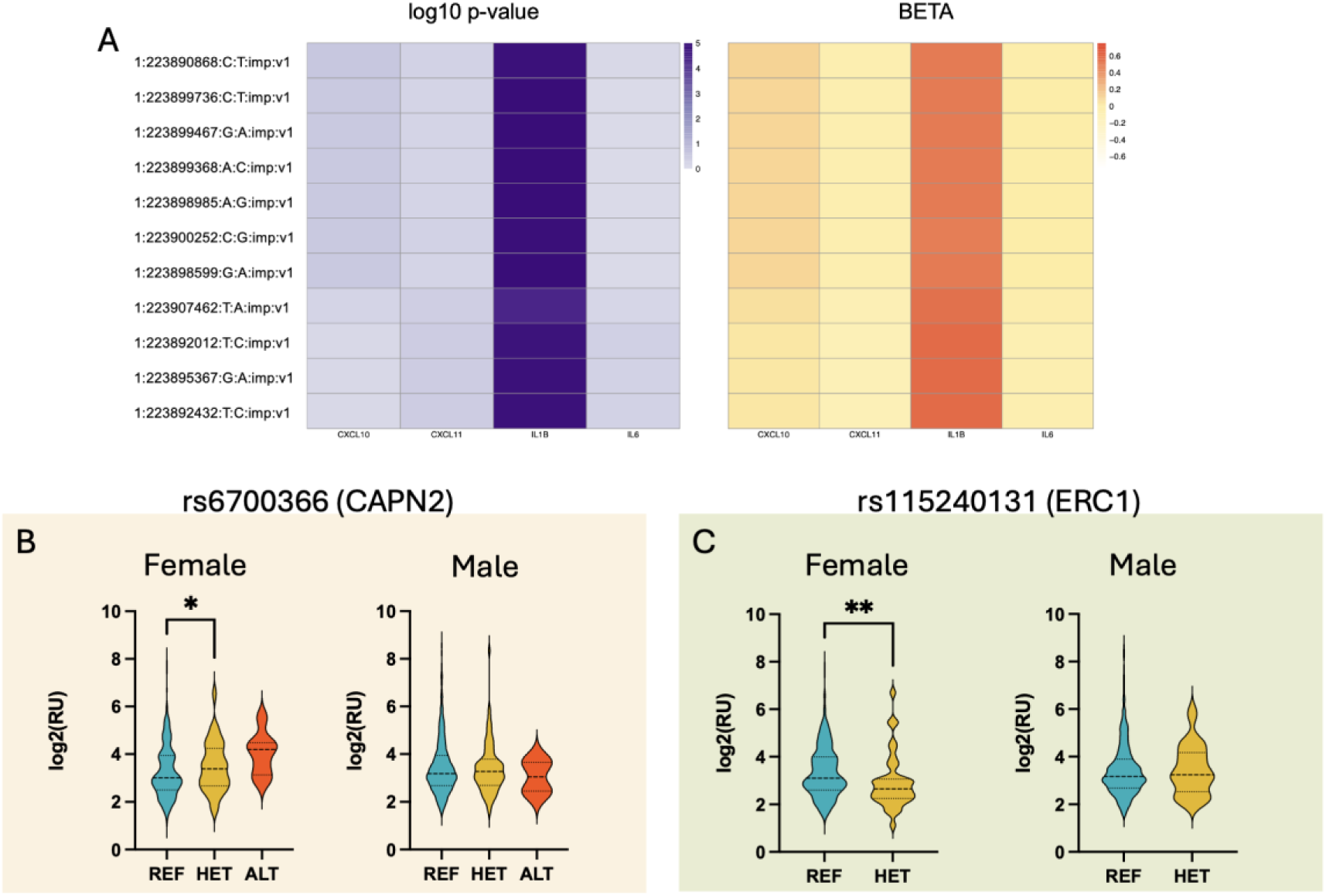
Plasma protein levels of NF-**к**B-related proteins. (A) Heatmap of proteomic AFR GWAS (pGWAS) results from UKBB identified a ancestry-specific association between variants near CAPN2 and IL-1B. (B,C) Violin plots summarizing sex-stratified plasma levels of NEMO protein in PMBB show increased levels in female individuals with CAPN2 variant, decreased levels in females with ERC1 variant, and no change in males.

The NF-**к**B signaling pathway is complex and context-specific, with alternative functions in distinct cell types. Furthermore, IL-1B represents both an activating ligand of the pathway and a downstream effector, and both of these are highly dependent on the third component of IKK complex, IKKg or NK-KB essential modulator (NEMO). We therefore measured plasma NEMO levels in the PET perfusion cohort using O-link CV II panel. Females, but not males, with rs6700366 variant had 12.3% increase in plasma NEMO levels (p<0.05, Figure 4B). rs115240131 also had sex-specific effects but in the opposite direction such that females with the variant had 14.4% lower plasma NEMO levels (p<0.01, Figure 4C). In general, these data support an association between rs6700366 and rs115240131 and altered NF-**к**B pathway signaling.

### Sex-based heterogeneity effects

Given the sex-differences identified in several of our analyses, we assessed for sex-based heterogeneity effects using GWAS in the AFR and EUR datasets separately and then meta-analyzed the sex-stratified results to assess differential effects by sex (Figure 1, inner tracks). Upon filtering for a heterogeneity p-value threshold of <5e-4, we identified 2,045 variants for the AFR population and 731 variants for the EUR population. We identified 2,771 unique variants spanning 95 distinct loci that exhibited evidence of heterogeneity based on having an I˄2 statistic > 40 between male and female participants. At this p-value threshold, we noted increased heterogeneity in the AFR population relative to the EUR population, based on the I˄2 values, which measure the proportion of variation due to heterogeneity in meta-analyzed studies (Supplemental Tables 8, 9).

## Discussion

In this study, we present the first unbiased GWAS for CMVD using perfusion PET flow reserve as the phenotype. Subsequent fine-mapping identified the rs115240131 near *ERC1*, and TWAS identified rs6700366 near *CAPN2* as putative risk-loci. Using coronary angiography data in distinct PMBB population, we confirmed these variants were associated with improved TIMI frame count, a measure of the coronary microvasculature, and showed no difference in obstructive CAD, supporting the relationship between these variants and CMVD rather than CAD (Figure 2). We next performed pathway analysis which converged on the NF-**к**B pathway and used proteomic data to show and association between dysregulation of NF-**к**B pathway and these variants (Figure 5).

Our study suggests that disrupting NF-KB signaling may be protective, perhaps by modifying cardiac anti-angiogenic signaling (e.g. through CXCL10 and CXCL11) to affect the coronary microvasculature. Interestingly, proteomic data pointed to distinct patterns of NF-**к**B signaling in individuals with rs115240131 versus those with rs6700366. Specifically, IL-1B was increased in individuals with rs6700366 in UKBB, but not those with rs115240131. NEMO was also differentially regulated between the pathway with increased NEMO in females with rs6700366 and decreased NEMO in females with rs115240131. The NF-KB pathway is known to have canonical and non-canonical arms in response to different stimuli and with distinct downstream effects. rs6700366 is associated with *CAPN2*, which regulates the non-canonical arm. In these individuals, we see an increase in IL-1B and NEMO, which suggests intact and even increased canonical NF-**к**B signaling. In contrast, rs115240131 may affect ERC1, a member of the canonical signaling pathway, and therefore we see no increase in IL-1B, and a decrease in ERC1’s effector partner NEMO. Canonical and non-canonical NF-**к**B signaling are extremely nuanced, complex, and context dependent. For example, IL1B is a well-established target of NF-**к**B, however, canonical signaling has been shown to also negatively regulate IL-1B in certain contexts.^36^ Prior studies have linked IL-1B to CMVD.^37^ Our studies confirm in an unbiased manner a role for dysregulation of NF-**к**B pathway in CMVD. They also further highlight the complex relationship between the NF-**к**B pathway and CMVD. There is clearly a need for additional research to better understand the respective roles of canonical and non-canonical NF-**к**B signaling, the cell types involved, and the link to altered coronary flow reserve.

Our study also found other interesting signals. *AADAC* was a genome-wide significant locus. *AADAC* has been shown to be increased in VSMCs derived from individuals with diabetes who do not have atherosclerosis, suggesting a potentially protective effect.^10^ *AADAC* may play an important role in VSMC phenotype switching which is important in lipid metabolism and deposition, but also equally important in SMC migration and contractility, which can affect the coronary microvasculature. TWAS identified a second interesting locus near *PLA2G5* that is associated with MBFR. *PLA2G5* is a member of the secretory phospholipase A2 family and catalyzes the hydrolysis of membrane phospholipids to release lysophospholipid and free fatty acids, including arachidonic acid (AA). It is expressed in immune cells and ischemic human cardiomyocytes,^38^ among others,^39^ and PLA2G5 SNP rs11573191 has been associated with premature CAD and hypertension in a Mexican Mestizo population.^40^ Together, these associations point to other genes and pathways that may be involved in regulating the coronary microvasculature.

In addition to genome-wide analyses, we also performed a genome-association study between established CAD loci and MBFR. Given the heterogeneity of individuals included in CAD GWAS studies, it is likely that these loci represent genes and pathways relevant to CMVD as well.^10^ Several interesting loci including KLF4 and FLT1 identified in prior GWAS were significantly associated with MBFR. Of note, KLF4 has emerged as a key regulator of smooth muscle cell (SMC) phenotype switching in females,^41^ which could potentially affect proper SMC functioning and microvascular vasodilation in sex-dependent manner. This is particularly relevant given sex-heterogeneity seen in CMVD.^5^

Indeed, our analysis reinforced results from prior gene association studies that identified sex-specific loci associated with CMVD.^10^ We found that CMVD is both sex-and ancestry-specific, with distinct loci identified from individuals of AFR and EUR genetic ancestry and I˄2 statistic > 40 between male and female participants. The balanced sample sizes for both male and female participants in our study further bolster the reliability and generalizability of these identified associations. These genomic data are further bolstered by the plasma proteomic data. For example, plasma NEMO levels were noted to be different by genotype in females, but not in males (Figure 4B). These data highlight the sex-heterogeneity seen in CMVD and the importance of including significant representation from males and females of diverse genetic ancestry.

Our study includes several limitations. First, our sample size is low for GWAS analysis. However, despite this limitation, we were able to leverage a continuous variable and downstream analyses. Second, we use perfusion PET studies that have been performed for clinical indications and using standard clinical protocols. This introduces some variability in software over time which may decrease the sensitivity of our analysis. This also leads to bias in including a relatively sick population as even those individuals without CMVD had high rates of cardiovascular risk factors and indications to undergo cardiac stress testing. Future genomic studies are needed to replicate these results in diverse populations and additional studies are needed in pre-clinical models to understand the role of NF-**к**B in different issues, cell types, and contexts.

In conclusion, we were able to use retrospective EHR data to identify novel loci for CMVD, prioritize existing IHD loci, and further support a role for NF-**к**B pathway in CMVD. This study highlights the potential of genomic studies to accelerate mechanistic knowledge in a critical area of cardiovascular research.

Supplemental Tables available for review at the following link: https://upenn.box.com/s/1i0c0d074kmb0mviu4vex8nwt6bhkn77

## Data Availability

All summary data will be made available, and the supplemental tables include full analyses results. For review of files larger than 75MB, a link is included in the cover letter.

## Acknowledgements

This work was supported by Alpha Phi Heart to Heart (MG, SSV), NHLBI R01HL175485 and BWF CAMS (MG), NHGRI T32HG000046 (RK, LG), NIH K12GM081259 (TC), Doris Duke Foundation (Award 2023-0224, MGL) and US Department of Veterans Affairs Biomedical Research and Development Award IK2-BX006551 (MGL). This article does not represent the views of the Department of Veterans Affairs or the US government.

We acknowledge the Penn Medicine BioBank (PMBB) for providing data and thank the patient-participants of Penn Medicine who consented to participate in this research program. We would also like to thank the Penn Medicine BioBank team and Regeneron Genetics Center for providing genetic variant data for analysis. The PMBB is approved under IRB protocol# 813913 and supported by Perelman School of Medicine at University of Pennsylvania, a gift from the Smilow family, and the National Center for Advancing Translational Sciences of the National Institutes of Health under CTSA award number UL1TR001878. A full list of contributors in included in Supplemental Note 1.

## References

1. Crea F, Camici PG and Bairey Merz CN. Coronary microvascular dysfunction: an update. Eur Heart J. 2014;35:1101–11.

2. Murthy VL, Naya M, Taqueti VR, Foster CR, Gaber M, Hainer J, Dorbala S, Blankstein R, Rimoldi O, Camici PG and Di Carli MF. Effects of sex on coronary microvascular dysfunction and cardiac outcomes. Circulation. 2014;129:2518–27.

3. Petersen JW, Johnson BD, Kip KE, Anderson RD, Handberg EM, Sharaf B, Mehta PK, Kelsey SF, Merz CN and Pepine CJ. TIMI frame count and adverse events in women with no obstructive coronary disease: a pilot study from the NHLBI-sponsored Women’s Ischemia Syndrome Evaluation (WISE). PloS one. 2014;9:e96630.

4. Sattar A, Mondragon, J. Clegg, S., Sheldon, M., Ahmed, B. Prevalence and Predictors of Conrary Microvascualr Dysfunction among Patients with Ischemia and Non-obstrucive CAD. J Am Coll Cardiol. 2016;67:1.

5. Bairey Merz CN, Pepine CJ, Walsh MN and Fleg JL. Ischemia and No Obstructive Coronary Artery Disease (INOCA): Developing Evidence-Based Therapies and Research Agenda for the Next Decade. Circulation. 2017;135:1075–1092.

6. Guerraty MA, Rao HS, Anjan VY, Szapary H, Mankoff DA, Pryma DA, Rader DJ and Dubroff JG. The role of resting myocardial blood flow and myocardial blood flow reserve as a predictor of major adverse cardiovascular outcomes. Plos One. 2020;15.

7. McPherson R and Tybjaerg-Hansen A. Genetics of Coronary Artery Disease. Circ Res. 2016;118:564–78.

8. Aragam KG, Jiang T, Goel A, Kanoni S, Wolford BN, Atri DS, Weeks EM, Wang M, Hindy G, Zhou W, Grace C, Roselli C, Marston NA, Kamanu FK, Surakka I, Venegas LM, Sherliker P, Koyama S, Ishigaki K, Asvold BO, Brown MR, Brumpton B, de Vries PS, Giannakopoulou O, Giardoglou P, Gudbjartsson DF, Guldener U, Haider SMI, Helgadottir A, Ibrahim M, Kastrati A, Kessler T, Kyriakou T, Konopka T, Li L, Ma L, Meitinger T, Mucha S, Munz M, Murgia F, Nielsen JB, Nothen MM, Pang S, Reinberger T, Schnitzler G, Smedley D, Thorleifsson G, von Scheidt M, Ulirsch JC, Biobank J, Epic CVD, Arnar DO, Burtt NP, Costanzo MC, Flannick J, Ito K, Jang DK, Kamatani Y, Khera AV, Komuro I, Kullo IJ, Lotta LA, Nelson CP, Roberts R, Thorgeirsson G, Thorsteinsdottir U, Webb TR, Baras A, Bjorkegren JLM, Boerwinkle E, Dedoussis G, Holm H, Hveem K, Melander O, Morrison AC, Orho-Melander M, Rallidis LS, Ruusalepp A, Sabatine MS, Stefansson K, Zalloua P, Ellinor PT, Farrall M, Danesh J, Ruff CT, Finucane HK, Hopewell JC, Clarke R, Gupta RM, Erdmann J, Samani NJ, Schunkert H, Watkins H, Willer CJ, Deloukas P, Kathiresan S, Butterworth AS and Consortium CAD. Discovery and systematic characterization of risk variants and genes for coronary artery disease in over a million participants. Nat Genet. 2022;54:1803–1815.

9. Schunkert H, Konig IR, Kathiresan S, Reilly MP, Assimes TL, Holm H, Preuss M, Stewart AF, Barbalic M, Gieger C, Absher D, Aherrahrou Z, Allayee H, Altshuler D, Anand SS, Andersen K, Anderson JL, Ardissino D, Ball SG, Balmforth AJ, Barnes TA, Becker DM, Becker LC, Berger K, Bis JC, Boekholdt SM, Boerwinkle E, Braund PS, Brown MJ, Burnett MS, Buysschaert I, Cardiogenics, Carlquist JF, Chen L, Cichon S, Codd V, Davies RW, Dedoussis G, Dehghan A, Demissie S, Devaney JM, Diemert P, Do R, Doering A, Eifert S, Mokhtari NE, Ellis SG, Elosua R, Engert JC, Epstein SE, de Faire U, Fischer M, Folsom AR, Freyer J, Gigante B, Girelli D, Gretarsdottir S, Gudnason V, Gulcher JR, Halperin E, Hammond N, Hazen SL, Hofman A, Horne BD, Illig T, Iribarren C, Jones GT, Jukema JW, Kaiser MA, Kaplan LM, Kastelein JJ, Khaw KT, Knowles JW, Kolovou G, Kong A, Laaksonen R, Lambrechts D, Leander K, Lettre G, Li M, Lieb W, Loley C, Lotery AJ, Mannucci PM, Maouche S, Martinelli N, McKeown PP, Meisinger C, Meitinger T, Melander O, Merlini PA, Mooser V, Morgan T, Muhleisen TW, Muhlestein JB, Munzel T, Musunuru K, Nahrstaedt J, Nelson CP, Nothen MM, Olivieri O, Patel RS, Patterson CC, Peters A, Peyvandi F, Qu L, Quyyumi AA, Rader DJ, Rallidis LS, Rice C, Rosendaal FR, Rubin D, Salomaa V, Sampietro ML, Sandhu MS, Schadt E, Schafer A, Schillert A, Schreiber S, Schrezenmeir J, Schwartz SM, Siscovick DS, Sivananthan M, Sivapalaratnam S, Smith A, Smith TB, Snoep JD, Soranzo N, Spertus JA, Stark K, Stirrups K, Stoll M, Tang WH, Tennstedt S, Thorgeirsson G, Thorleifsson G, Tomaszewski M, Uitterlinden AG, van Rij AM, Voight BF, Wareham NJ, Wells GA, Wichmann HE, Wild PS, Willenborg C, Witteman JC, Wright BJ, Ye S, Zeller T, Ziegler A, Cambien F, Goodall AH, Cupples LA, Quertermous T, Marz W, Hengstenberg C, Blankenberg S, Ouwehand WH, Hall AS, Deloukas P, Thompson JR, Stefansson K, Roberts R, Thorsteinsdottir U, O’Donnell CJ, McPherson R, Erdmann J, Consortium CA and Samani NJ. Large-scale association analysis identifies 13 new susceptibility loci for coronary artery disease. Nat Genet. 2011;43:333–8.

10. Wayne N, Singamneni VS, Venkatesh R, Cherlin T, Verma SS and Guerraty MA. Genetic Insights Into Coronary Microvascular Disease. Microcirculation. 2025;32:e12896.

11. Verma A, Damrauer SM, Naseer N, Weaver J, Kripke CM, Guare L, Sirugo G, Kember RL, Drivas TG, Dudek SM, Bradford Y, Lucas A, Judy R, Verma SS, Meagher E, Nathanson KL, Feldman M, Ritchie MD, Rader DJ and For The Penn Medicine B. The Penn Medicine BioBank: Towards a Genomics-Enabled Learning Healthcare System to Accelerate Precision Medicine in a Diverse Population. J Pers Med. 2022;12.

12. Genomes Project C, Auton A, Brooks LD, Durbin RM, Garrison EP, Kang HM, Korbel JO, Marchini JL, McCarthy S, McVean GA and Abecasis GR. A global reference for human genetic variation. Nature. 2015;526:68–74.

13. Wayne N, Wu Q, Moore SC, Ferrari VA, Metzler SD and Guerraty MA. Multimodality assessment of the coronary microvasculature with TIMI frame count versus perfusion PET highlights coronary changes characteristic of coronary microvascular disease. Frontiers in Cardiovascular Medicine. 2024;11.

14. Gibson CM, Cannon CP, Daley WL, Dodge JT, Alexander B, Marble SJ, McCabe CH, Raymond L, Fortin T, Poole WK and Braunwald E. TIMI frame count: A quantitative method of assessing coronary artery flow. Circulation. 1996;93:879–888.

15. Mbatchou J, Barnard L, Backman J, Marcketta A, Kosmicki JA, Ziyatdinov A, Benner C, O’Dushlaine C, Barber M, Boutkov B, Habegger L, Ferreira M, Baras A, Reid J, Abecasis G, Maxwell E and Marchini J. Computationally efficient whole-genome regression for quantitative and binary traits. Nat Genet. 2021;53:1097–1103.

16. Chang CC, Chow CC, Tellier LC, Vattikuti S, Purcell SM and Lee JJ. Second-generation PLINK: rising to the challenge of larger and richer datasets. Gigascience. 2015;4:7.

17. Wang G, Sarkar A, Carbonetto P and Stephens M. A simple new approach to variable selection in regression, with application to genetic fine mapping. J R Stat Soc Series B Stat Methodol. 2020;82:1273–1300.

18. Barbeira AN, Dickinson SP, Bonazzola R, Zheng J, Wheeler HE, Torres JM, Torstenson ES, Shah KP, Garcia T, Edwards TL, Stahl EA, Huckins LM, Consortium GT, Nicolae DL, Cox NJ and Im HK. Exploring the phenotypic consequences of tissue specific gene expression variation inferred from GWAS summary statistics. Nat Commun. 2018;9:1825.

19. Cordero P, Parikh VN, Chin ET, Erbilgin A, Gloudemans MJ, Shang C, Huang Y, Chang AC, Smith KS, Dewey F, Zaleta K, Morley M, Brandimarto J, Glazer N, Waggott D, Pavlovic A, Zhao M, Moravec CS, Tang WHW, Skreen J, Malloy C, Hannenhalli S, Li H, Ritter S, Li M, Bernstein D, Connolly A, Hakonarson H, Lusis AJ, Margulies KB, Depaoli-Roach AA, Montgomery SB, Wheeler MT, Cappola T and Ashley EA. Pathologic gene network rewiring implicates PPP1R3A as a central regulator in pressure overload heart failure. Nat Commun. 2019;10:2760.

20. Gamazon ER, Wheeler HE, Shah KP, Mozaffari SV, Aquino-Michaels K, Carroll RJ, Eyler AE, Denny JC, Consortium GT, Nicolae DL, Cox NJ and Im HK. A gene-based association method for mapping traits using reference transcriptome data. Nat Genet. 2015;47:1091–8.

21. Liu Y, Morley M, Brandimarto J, Hannenhalli S, Hu Y, Ashley EA, Tang WH, Moravec CS, Margulies KB, Cappola TP, Li M and consortium MA. RNA-Seq identifies novel myocardial gene expression signatures of heart failure. Genomics. 2015;105:83–9.

22. Subramanian A, Tamayo P, Mootha VK, Mukherjee S, Ebert BL, Gillette MA, Paulovich A, Pomeroy SL, Golub TR, Lander ES and Mesirov JP. Gene set enrichment analysis: a knowledge-based approach for interpreting genome-wide expression profiles. Proc Natl Acad Sci U S A. 2005;102:15545–50.

23. Sun BB, Chiou J, Traylor M, Benner C, Hsu YH, Richardson TG, Surendran P, Mahajan A, Robins C, Vasquez-Grinnell SG, Hou L, Kvikstad EM, Burren OS, Davitte J, Ferber KL, Gillies CE, Hedman AK, Hu S, Lin T, Mikkilineni R, Pendergrass RK, Pickering C, Prins B, Baird D, Chen CY, Ward LD, Deaton AM, Welsh S, Willis CM, Lehner N, Arnold M, Worheide MA, Suhre K, Kastenmuller G, Sethi A, Cule M, Raj A, Alnylam Human G, AstraZeneca Genomics I, Biogen Biobank T, Bristol Myers S, Genentech Human G, GlaxoSmithKline Genomic S, Pfizer Integrative B, Population Analytics of Janssen Data S, Regeneron Genetics C, Burkitt-Gray L, Melamud E, Black MH, Fauman EB, Howson JMM, Kang HM, McCarthy MI, Nioi P, Petrovski S, Scott RA, Smith EN, Szalma S, Waterworth DM, Mitnaul LJ, Szustakowski JD, Gibson BW, Miller MR and Whelan CD. Plasma proteomic associations with genetics and health in the UK Biobank. Nature. 2023;622:329–338.

24. May D, Gilon D, Djonov V, Itin A, Lazarus A, Gordon O, Rosenberger C and Keshet E. Transgenic system for conditional induction and rescue of chronic myocardial hibernation provides insights into genomic programs of hibernation. Proc Natl Acad Sci U S A. 2008;105:282–7.

25. Haskins RM, Nguyen AT, Alencar GF, Billaud M, Kelly-Goss MR, Good ME, Bottermann K, Klibanov AL, French BA, Harris TE, Peirce SM, Isakson BE and Owens GK. Klf4 has an unexpected protective role in perivascular cells within the microvasculature. Am J Physiol Heart Circ Physiol. 2018;315:H402–H414.

26. SenthilKumar G, Katunaric B, Zirgibel Z, Lindemer B, Jaramillo-Torres MJ, Bordas-Murphy H, Schulz ME, Pearson PJ and Freed JK. Necessary Role of Acute Ceramide Formation in The Human Microvascular Endothelium During Health and Disease. bioRxiv. 2023.

27. Toyohara T, Roudnicky F, Florido MHC, Nakano T, Yu H, Katsuki S, Lee M, Meissner T, Friesen M, Davidow LS, Ptaszek L, Abe T, Rubin LL, Pereira AC, Aikawa M and Cowan CA. Patient hiPSCs Identify Vascular Smooth Muscle Arylacetamide Deacetylase as Protective against Atherosclerosis. Cell Stem Cell. 2020;27:147–157 e7.

28. Fassett J, Xu X, Kwak D, Zhu G, Fassett EK, Zhang P, Wang H, Mayer B, Bache RJ and Chen Y. Adenosine kinase attenuates cardiomyocyte microtubule stabilization and protects against pressure overload-induced hypertrophy and LV dysfunction. J Mol Cell Cardiol. 2019;130:49–58.

29. Aguilar-Salinas CA, Tusie-Luna T and Pajukanta P. Genetic and environmental determinants of the susceptibility of Amerindian derived populations for having hypertriglyceridemia. Metabolism. 2014;63:887–94.

30. Khounphinith E, Yin RX, Cao XL, Huang F, Wu JZ and Li H. TIMD4 rs6882076 SNP Is Associated with Decreased Levels of Triglycerides and the Risk of Coronary Heart Disease and Ischemic Stroke. Int J Med Sci. 2019;16:864–871.

31. Carss KJ, Baranowska AA, Armisen J, Webb TR, Hamby SE, Premawardhana D, Al-Hussaini A, Wood A, Wang Q, Deevi SVV, Vitsios D, Lewis SH, Kotecha D, Bouatia-Naji N, Hesselson S, Iismaa SE, Tarr I, McGrath-Cadell L, Muller DW, Dunwoodie SL, Fatkin D, Graham RM, Giannoulatou E, Samani NJ, Petrovski S, Haefliger C and Adlam D. Spontaneous Coronary Artery Dissection: Insights on Rare Genetic Variation From Genome Sequencing. Circ Genom Precis Med. 2020;13:e003030.

32. Wayne N, Wu Q, Moore SC, Ferrari VA, Metzler SD and Guerraty MA. Multimodality assessment of the coronary microvasculature with TIMI frame count versus perfusion PET highlights coronary changes characteristic of coronary microvascular disease. Front Cardiovasc Med. 2024;11:1395036.

33. Ducut Sigala JL, Bottero V, Young DB, Shevchenko A, Mercurio F and Verma IM. Activation of transcription factor NF-kappaB requires ELKS, an IkappaB kinase regulatory subunit. Science. 2004;304:1963–7.

34. Shumway SD, Maki M and Miyamoto S. The PEST domain of IkappaBalpha is necessary and sufficient for in vitro degradation by mu-calpain. J Biol Chem. 1999;274:30874–81.

35. Tabula Sapiens C, Jones RC, Karkanias J, Krasnow MA, Pisco AO, Quake SR, Salzman J, Yosef N, Bulthaup B, Brown P, Harper W, Hemenez M, Ponnusamy R, Salehi A, Sanagavarapu BA, Spallino E, Aaron KA, Concepcion W, Gardner JM, Kelly B, Neidlinger N, Wang Z, Crasta S, Kolluru S, Morri M, Tan SY, Travaglini KJ, Xu C, Alcantara-Hernandez M, Almanzar N, Antony J, Beyersdorf B, Burhan D, Calcuttawala K, Carter MM, Chan CKF, Chang CA, Chang S, Colville A, Culver RN, Cvijovic I, D’Amato G, Ezran C, Galdos FX, Gillich A, Goodyer WR, Hang Y, Hayashi A, Houshdaran S, Huang X, Irwin JC, Jang S, Juanico JV, Kershner AM, Kim S, Kiss B, Kong W, Kumar ME, Kuo AH, Li B, Loeb GB, Lu WJ, Mantri S, Markovic M, McAlpine PL, de Morree A, Mrouj K, Mukherjee S, Muser T, Neuhofer P, Nguyen TD, Perez K, Puluca N, Qi Z, Rao P, Raquer-McKay H, Schaum N, Scott B, Seddighzadeh B, Segal J, Sen S, Sikandar S, Spencer SP, Steffes LC, Subramaniam VR, Swarup A, Swift M, Van Treuren W, Trimm E, Veizades S, Vijayakumar S, Vo KC, Vorperian SK, Wang W, Weinstein HNW, Winkler J, Wu TTH, Xie J, Yung AR, Zhang Y, Detweiler AM, Mekonen H, Neff NF, Sit RV, Tan M, Yan J, Bean GR, Charu V, Forgo E, Martin BA, Ozawa MG, Silva O, Toland A, Vemuri VNP, Afik S, Awayan K, Botvinnik OB, Byrne A, Chen M, Dehghannasiri R, Gayoso A, Granados AA, Li Q, Mahmoudabadi G, McGeever A, Olivieri JE, Park M, Ravikumar N, Stanley G, Tan W, Tarashansky AJ, Vanheusden R, Wang P, Wang S, Xing G, Dethlefsen L, Ezran C, Gillich A, Hang Y, Ho PY, Irwin JC, Jang S, Leylek R, Liu S, Maltzman JS, Metzger RJ, Phansalkar R, Sasagawa K, Sinha R, Song H, Swarup A, Trimm E, Veizades S, Wang B, Beachy PA, Clarke MF, Giudice LC, Huang FW, Huang KC, Idoyaga J, Kim SK, Kuo CS, Nguyen P, Rando TA, Red-Horse K, Reiter J, Relman DA, Sonnenburg JL, Wu A, Wu SM and Wyss-Coray T. The Tabula Sapiens: A multiple-organ, single-cell transcriptomic atlas of humans. Science. 2022;376:eabl4896.

36. Greten FR, Arkan MC, Bollrath J, Hsu LC, Goode J, Miething C, Goktuna SI, Neuenhahn M, Fierer J, Paxian S, Van Rooijen N, Xu Y, O’Cain T, Jaffee BB, Busch DH, Duyster J, Schmid RM, Eckmann L and Karin M. NF-kappaB is a negative regulator of IL-1beta secretion as revealed by genetic and pharmacological inhibition of IKKbeta. Cell. 2007;130:918–31.

37. Taqueti VR, Shah AM, Everett BM, Pradhan AD, Piazza G, Bibbo C, Hainer J, Morgan V, Carolina do AHdSA, Skali H, Blankstein R, Dorbala S, Goldhaber SZ, Le May MR, Chow BJW, deKemp RA, Hage FG, Beanlands RS, Libby P, Glynn RJ, Solomon SD, Ridker PM and Di Carli MF. Coronary Flow Reserve, Inflammation, and Myocardial Strain: The CIRT-CFR Trial. JACC Basic Transl Sci. 2023;8:141–151.

38. Ishikawa Y, Komiyama K, Masuda S, Murakami M, Akasaka Y, Ito K, Akishima-Fukasawa Y, Kimura M, Fujimoto A, Kudo I and Ishii T. Expression of type V secretory phospholipase A in myocardial remodelling after infarction. Histopathology. 2005;47:257–67.

39. Samuchiwal SK and Balestrieri B. Harmful and protective roles of group V phospholipase A(2): Current perspectives and future directions. Biochim Biophys Acta Mol Cell Biol Lipids. 2019;1864:819–826.

40. Vargas-Alarcon G, Posadas-Romero C, Villarreal-Molina T, Alvarez-Leon E, Angeles-Martinez J, Soto ME, Monroy-Munoz I, Juarez JG, Sanchez-Ramirez CJ, Ramirez-Bello J, Ramirez-Fuentes S, Fragoso JM and Rodriguez-Perez JM. The (G>A) rs11573191 polymorphism of PLA2G5 gene is associated with premature coronary artery disease in the Mexican Mestizo population: the genetics of atherosclerotic disease Mexican study. Biomed Res Int. 2014;2014:931361.

41. Hartman RJG, Owsiany K, Ma L, Koplev S, Hao K, Slenders L, Civelek M, Mokry M, Kovacic JC, Pasterkamp G, Owens G, Bjorkegren JLM and den Ruijter HM. Sex-Stratified Gene Regulatory Networks Reveal Female Key Driver Genes of Atherosclerosis Involved in Smooth Muscle Cell Phenotype Switching. Circulation. 2021;143:713–726.

